# Roles of childhood maltreatment, resilience and sleep disturbances on quality of life in chronic pain

**DOI:** 10.64898/2025.12.02.25341513

**Authors:** Rebecca Peters, Fabiènne Schinke, Sylvia M. Gustin, Inga Schalinski, Yann Quidé

## Abstract

**Background:** Around one in five persons globally reports experiencing chronic pain. Chronic pain can lead to sleep disturbances, reduced resilience to chronic stressors, and quality of life. Exposure to childhood maltreatment is a risk factor for chronic pain that also reduces resilience abilities. However, the relationship between childhood maltreatment, resilience and sleep disturbance on quality of life in people with chronic pain remain poorly understood.

**Methods:** Two hundred and forty-one participants were included in this study, including 157 people with and 84 without chronic pain (controls). All participants responded to an online survey that included measures of childhood maltreatment, resilience, sleep disturbances and quality of life. A moderated serial mediation model tested how resilience and sleep disturbances mediate the relationship between group and quality of life, and how the severity of childhood maltreatment moderates the group difference in resilience.

**Results:** Participants with chronic pain reported significantly lower quality of life than controls. This group difference in quality of life was significantly mediated by the level of resilience and sleep disturbance when people reported being exposed to low and average, but not high levels of childhood maltreatment.

**Conclusions:** The study found that exposure to childhood maltreatment reduced resilience abilities leading to more severe sleep problems and lower quality of life in participants with chronic pain. These findings suggest that interventions targeting childhood maltreatment and resilience may be beneficial to improve quality of life, through increased sleep quality, in individuals with chronic pain.

## Introduction

Chronic pain is experienced by approximately 20-30% of the global population (Cohen, Vase et al. 2021, Turk and Patel 2022). Many individuals with chronic pain feel misunderstood, the legitimacy of their pain being questioned, especially by healthcare professionals (Nicola, Correia et al. 2021). The large treatment gap for chronic pain (Kieselbach, Schiltenwolf et al. 2016) increases the economic burden on the healthcare system (Breivik, Collett et al. 2006), and risks for the development of psychopathologies (Arnow, Hunkeler et al. 2006, Burke, Mathias et al. 2015). Chronic pain has negative impacts on mood, sleep and quality of life (Dysvik, Lindstrøm et al. 2004, Yazdi-Ravandi, Taslimi et al. 2013, Hadi, McHugh et al. 2019, Wörz, Horlemann et al. 2022). Other risk factors for chronic pain, such as exposure to childhood maltreatment, can further reduce resilience (Davis, Luecken et al. 2005, Burke, Finn et al. 2017), increasing pain-related sleep problems and poorer quality of life. However, the exact relationship between childhood maltreatment, resilience, sleep problems and quality of life within the same group of individuals experiencing, or not, chronic pain remains unclear, highlighting an important gap in the literature.

Quality of life is a multi-faceted construct that considers both negative and positive aspects of different areas of life (Teoli and Bhardwaj 2024). In particular, reduced health-related quality of life, including physical, psychological, and social independent dimensions (Karimi and Brazier 2016) is consistently reported in chronic pain (Hunfeld, Perquin et al. 2001, Dysvik, Lindstrøm et al. 2004, Phyo, Freak-Poli et al. 2020). In separate studies, chronic pain, but also poor sleep quality, and poor resilience skills have been associated with reduced quality of life. Many factors can impact quality of life with chronic pain being one of the most severe (Hunfeld, Perquin et al. 2001, Dysvik, Lindstrøm et al. 2004), and quality of sleep (Reimer and Flemons 2003, Carpi, Cianfarani et al. 2022), childhood maltreatment (Greger, Myhre et al. 2016) and resilience skills (Chng, Yeo et al. 2023) being other significant ones. In particular, poor sleep quality, experienced in 50-60% of people with chronic pain, negatively influences their quality of life (Veldhuijzen, Greenspan et al. 2008, Purabdollah, Lakdizaji et al. 2015, Carpi, Cianfarani et al. 2022), resulting in increased and longer-lasting pain intensity in people with chronic pain (Husak and Bair 2020).

Beneath sleep disturbances, resilience and the experience of childhood maltreatment (Davis, Luecken et al. 2005, Weber, Jud et al. 2016, Morete, Solano et al. 2018, Chng, Yeo et al. 2023) are factors that can contribute to the impacts of chronic pain on quality of life. Patients with chronic pain are more likely to report a history of childhood maltreatment compared to people who do not experience chronic pain (Davis, Luecken et al. 2005, Edwards, Dworkin et al. 2016, Burke, Finn et al. 2017, Studer, Stewart et al. 2017, Beal, Kashikar-Zuck et al. 2020). In addition, chronic pain is also associated with low resilience to stress (Morete, Solano et al. 2018, Kascakova, Furstova et al. 2022), and independently of experiencing chronic pain, childhood maltreatment is associated with poorer resilience skills (Ding, Han et al. 2017, Fritz, de Graaff et al. 2018, Park, Lee et al. 2023). It is therefore possible that the severity of childhood maltreatment exposure may interact with chronic pain to further reduce resilience, which in turn can lead to poor sleep quality and lower quality of life. However, the exact relationship existing between these factors has not been studied at the same time.

This study aims to fill this gap and identify how resilience skills and sleep quality account for (mediate) the difference in quality of life observed between participants experiencing, or not, chronic pain. Lower levels of resilience associated with chronic pain are expected to directly impact sleep quality (Liu, Liu et al. 2016, Cai, Wang et al. 2021, Arora, Grey et al. 2022, Chng, Yeo et al. 2023), which in turn will affect quality of life. In addition, the group difference on levels of resilience is expected to further depend (moderated by) on the severity of childhood maltreatment exposure.

## Method

### Participants

Cross-sectional data from 415 individuals aged 18 years or older, were collected in Germany between April and June 2024 using the online platform SoSci-Survey (Leiner 2024), distributed through social media (Instagram, Facebook). A final convenience sample of 241 individuals included 157 people reporting experiencing chronic pain and 84 people who did not (controls; see details Figure 1). This study was approved by the Ethics Committee of the University of the Bundeswehr Munich (EK UniBw M 24-20). Participants were informed about the study in writing and provided informed consent online prior to participation. Respondents were given the option ‘I don’t want to answer/I can’t answer’ to all questions. Participants were asked if the persistence of their pain was of at least three months (Treede, Rief et al. 2015). If the respondent answered “No” all subsequent pain-related questions were blanked out. Conversely, if the respondent answered “Yes” subsequent pain-related questions were presented (see details below). Additionally, respondents were queried about their use of painkillers to treat their condition.

**Figure 1.**
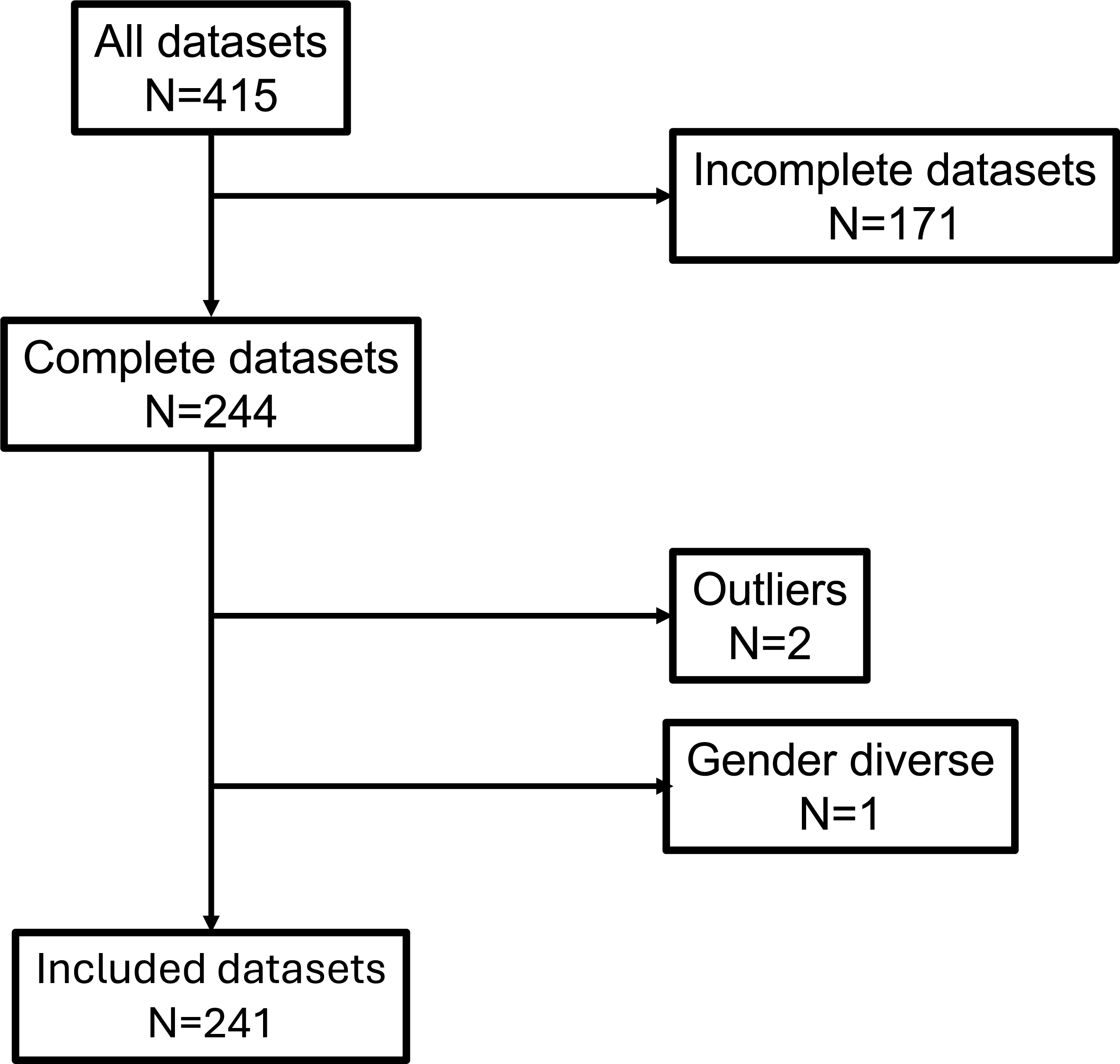
Flow diagram illustrating the inclusion of participants in the final analyses.

### Sociodemographic Data

Sociodemographic data included gender (male, female, non-binary), age, level of education, occupation, relationship status and health-related questions. The survey inquired about the presence of both psychological and physical illnesses. When respondents indicated the presence of a psychological or physical illness, they were asked to provide more detailed information about the specific condition using predefined responses such as “depression” or “asthma.”

The assessment of chronic pain was conducted using a set of standardized screening questions that have been frequently applied in the context of epidemiological research. Participants were first asked whether they had experienced continuous pain for a period of more than three months, reflecting the commonly used temporal criterion for chronic pain. In instances where this criterion was met, respondents were further invited to indicate whether they had received a medical pain diagnosis, to specify the type of chronic pain condition, to report the year of diagnosis, and to provide information on their use of pain medication. The sociodemographic dataset was constructed with specific reference to the presence of a chronic pain condition and the type of medication used.

Further pain-related information was collected for descriptive purposes, including ratings of current pain intensity and pain intensity during the preceding week (both assessed on 0–10 Numeric Rating Scales). In addition, validated questionnaires were used, such as the Pain Disability Index (Dillmann, Nilges et al. 2011), the Brief Pain Inventory (Radbruch, Loick et al. 1999) and the Tampa Scale of Kinesiophobia (Pagels, Lüdtke et al. 2022). However, for the statistical analyses, only the binary indicator of chronic pain was included. It is important to note that all additional pain-related measures were recorded; however, these were not incorporated into the analytical model.

### Quality of life

The Short-Form 36 Health Questionnaire (SF-36) was used to measure quality of life (Bullinger, Kirchberger et al. 1995, Morfeld, Kirchberger et al. 2011). This 36-items questionnaire measures different aspects of health-related quality of life, including vitality physical functioning, physical pain, general health perception, physical role functioning, emotional role functioning, social role functioning, psychological well-being and subjective change in health over the past year. An index of overall quality of life was derived as the total score to the SF-36, according to scoring guidelines. The questionnaire has been validated in different samples and shows a high internal consistency (Morfeld, Kirchberger et al. 2011). Previous chronic pain studies reported a moderate to high internal consistency with .61 < α < .93 (Gerbershagen, Lindena et al. 2002).

### Sleep Disturbances

The Patient-Reported Outcomes Measurement Information System (PROMIS) Sleep Disturbance 8b Questionnaire (PROMIS-SD8b) is an 8-items questionnaire used to measure sleep disturbances (Yu, Buysse et al. 2011). This five-point Likert scale (1 = “Not at all”, 5 = “Very Much”) record subjective ratings of sleep quality and sleep disturbance over the past 7 days. The PROMIS-SD8b total score (score range 8 – 40) was rescaled according to official guidelines and used as an index of the severity of sleep disturbances. The validation of the original English version of the questionnaire showed a high internal consistency of .89 < α < .96 (Buysse, Yu et al. 2010, Yu, Buysse et al. 2012).

### Childhood Maltreatment

The short form of the Childhood Trauma Questionnaire (CTQ) was used to assess the severity of childhood maltreatment exposure (Bernstein, Stein et al. 2003, Bader, Hänny et al. 2009). This 28-items questionnaire measures the severity of emotional, physical and sexual abuse, as well as emotional and physical neglect experiences in childhood and adolescence, using a 5-point Likert scale (1 = “Never true”, 5 = “Very often true”). The CTQ total score (based on 25 items; score range 25 – 125) was used as an index of the overall severity of childhood maltreatment. The three minimization/denial validity items are used separately to detect underreporting of trauma, but do not contribute to the calculation of the total score. The German version of the CTQ has high validity and reliability, with a Cronbach’s alpha, as an indicator of internal consistency, for the subscales between α = .74 and α = .94 (Bader, Hänny et al. 2009).

### Resilience

The Brief Resilience Scale (BRS) was used to assess the resilience of the respondents (Smith, Dalen et al. 2008, Chmitorz, Wenzel et al. 2018). The BRS consists of six items measuring the respondent’s general ability to recover after experiencing a crisis. The questionnaire is based on a five-point Likert scale (1 = “Strongly disagree”, 5 = “Strongly agree”). The BRS total score was used as an index of resilience (score range 6 – 30). In a validation study of the German version of the BRS, the scale showed good reliability with a Cronbach’s alpha of α = .85 (Chmitorz, Wenzel et al. 2018).

### Statistical Analysis

All statistical analyses were performed using IBM SPSS Statistics (Version 29.0.1.0). Incomplete datasets and those with missing values on the questionnaires of interest (CTQ, BRS, PROMIS-SD8b, and SF-36) were excluded (*N* = 171). The dataset was then examined for outliers using Mahalanobis distance, Cook’s distance, and leverage values. Datasets identified as outliers in at least two of the three tests were excluded (*N* = 2). The Kolmogorov-Smirnoff test indicated that no variables of interest, but the BRS total score, were normally distributed. Therefore, Mann-Whitney U tests were used to compare groups on levels of these variables, and an independent t-test was used to compare the BRS total scores between the groups.

The moderated mediation model was tested using the model 83 in PROCESS macro (v4.2) for SPSS (Hayes 2022). Group (chronic pain vs controls) was entered as the independent variable, quality of life (SF-36 total score) was the dependent variable, resilience (BRS total score) and sleep disturbances (PROMIS-SD8b total score) were the mediators, and the severity of childhood maltreatment (CTQ total score) was used as the moderator of the group difference in levels of resilience. A robust bootstrapping method was used for the mediation analysis, to account for the non-normal distribution of the variables of interest (Davison and Hinkley 1997). In case of significant association between the group-by-maltreatment interaction and resilience scores, moderation analysis was performed to determine the relationship between chronic pain and resilience at low (−1SD), average, and high (+1SD) levels of childhood maltreatment exposure (Cohen, Cohen et al. 2003). The HC3 (Davidson-MacKinnon) method was used to account for possible issues related to heteroskedasticity (Hayes and Cai 2007). Power analysis using G*Power v3.1.9.6 (Faul, Erdfelder et al. 2009) indicated that a sample size of 138 participants was necessary to have 95% power to detect a medium effect (*f^2^*= 0.15) at α = .05 in a serial mediation analysis with five predictors of the dependent variable (quality of life), including the independent variable (group), the mediators (resilience and sleep disturbance), and two covariates (age and gender).

## Results

### Sample characteristics

Sociodemographic and clinical characteristics of the sample studied are provided in Table 1 and Table 2. The chronic pain group was significantly older than the control group, but groups did not differ from each other regarding gender distribution, occupation and education (see Table 1). Regarding the variables of interest for this study, the chronic pain group reported significant higher levels of childhood maltreatment, lower resilience levels, greater sleep disturbances and lower quality of life compared to the control group (see Table 1). The chronic pain group reported being more often diagnosed with psychological or physical health conditions than the control group (see Table 2).

**Table 1.** Sociodemographic characteristics of the studied sample.

|  |  | Healthy Controls (N=84) |  | Chronic Pain (N=157) |  | Statistics |  |  |
| --- | --- | --- | --- | --- | --- | --- | --- | --- |
|  |  | Median | SD | Median | SD | Mann-Whitney U | z-statistics | p-values |
| Age, in years |  | 30.00 | 13.68 | 38.00 | 12.53 | **8488.50** | **3.675** | **<0.001** |
|  |  | N | % | N | % | X<sup>2</sup> | df | p-values |
| Gender (Females) |  | 50 | 60% | 142 | 90% | **32.302** | **1** | **<0.001** |
| Education |  |  |  |  |  | 4.846 | 5 | 0.435 |
|  | Ongoing | 5 | 6% | 13 | 8% |  |  |  |
|  | Secondary school leaving certificate (Hauptschulabschluss) | 27 | 32% | 52 | 33% |  |  |  |
|  | Intermediate secondary school leaving certificate (Realschulabschluss) | 12 | 14% | 28 | 18% |  |  |  |
|  | University entrance qualification (Fachhochschulreife) | 40 | 48% | 59 | 38% |  |  |  |
|  | A-Level (Abitur) | 0 | 0% | 1 | 0% |  |  |  |
|  | No information | 0 | 0% | 4 | 3% |  |  |  |
| Relationship status |  |  |  |  |  | 6.539 | 6 | 0.366 |
|  | Single | 19 | 23% | 32 | 20% |  |  |  |
|  | In a relationship | 33 | 39% | 47 | 30% |  |  |  |
|  | Defacto | 0 | 0% | 1 | 1% |  |  |  |
|  | Married | 29 | 35% | 65 | 41% |  |  |  |
|  | Divorced | 1 | 1% | 7 | 5% |  |  |  |
|  | Widowed | 2 | 2% | 2 | 1% |  |  |  |
|  | Other | 0 | 0% | 3 | 2% |  |  |  |
| Occupation |  |  |  |  |  | **26.270** | **7** | **<0.001** |
|  | Pupil | 0 | 0% | 2 | 1% |  |  |  |
|  | Student | 15 | 18% | 10 | 6% |  |  |  |
|  | Salaried employee | 45 | 54% | 83 | 53% |  |  |  |
|  | Civil servant | 12 | 14% | 7 | 4% |  |  |  |
|  | Self-employed | 4 | 5% | 6 | 4% |  |  |  |
|  | Unemployed | 0 | 0% | 5 | 3% |  |  |  |
|  | Pensioner | 5 | 6% | 30 | 19% |  |  |  |
|  | Other | 3 | 4% | 14 | 9% |  |  |  |

SD: standard deviation; df: degrees of freedom

**Table 2.**
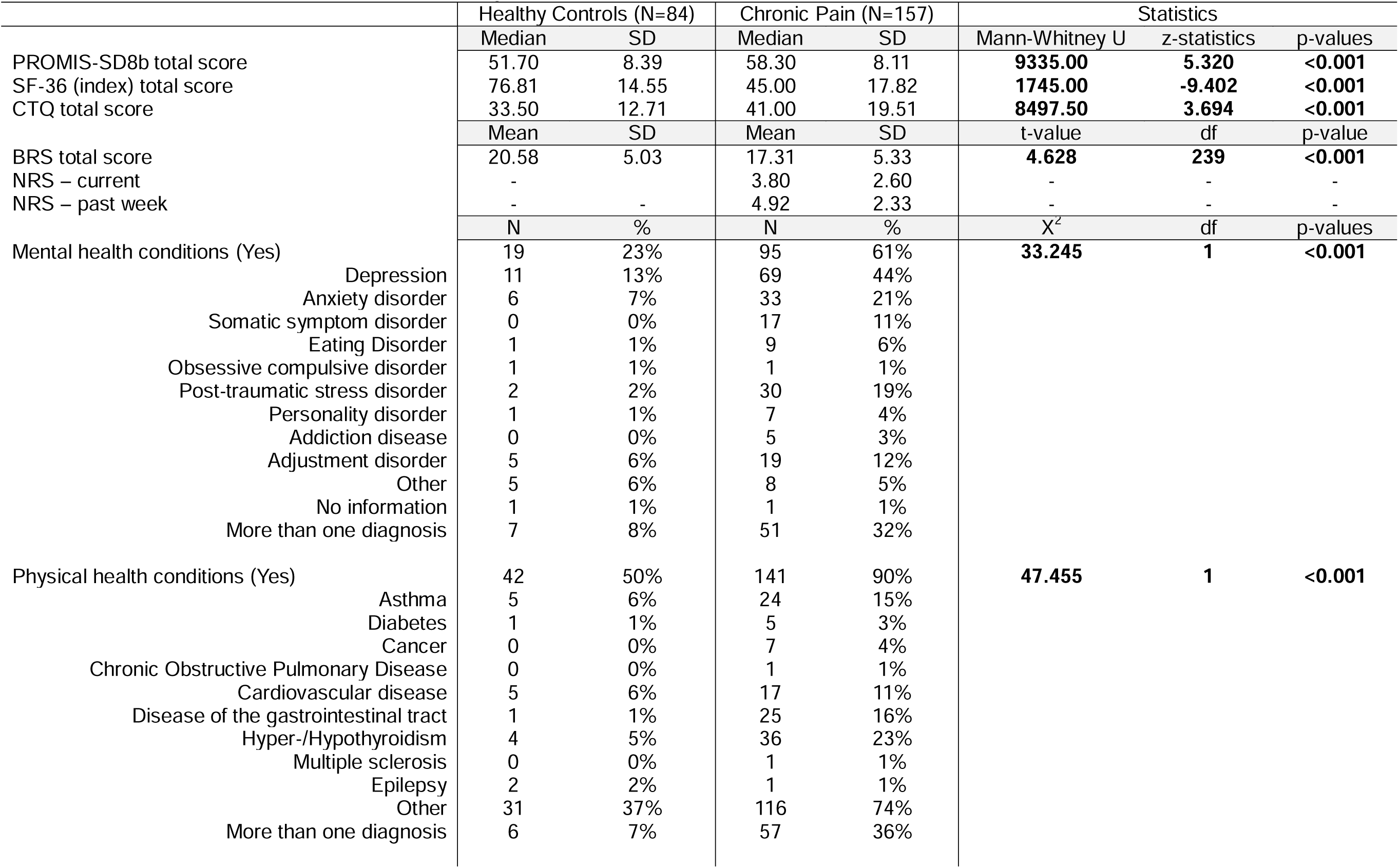
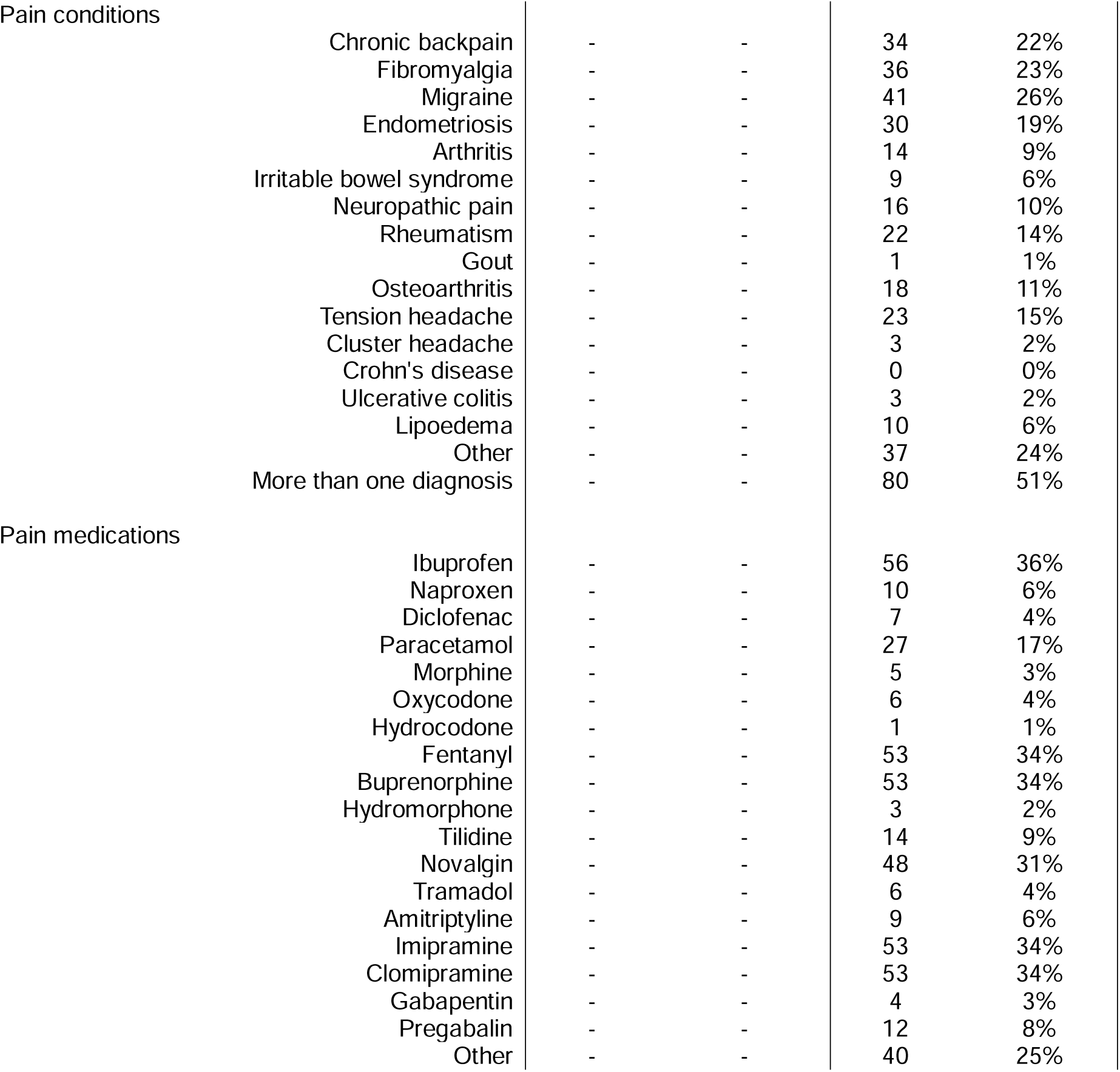
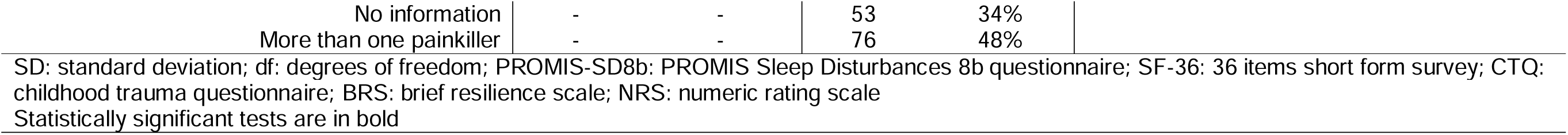
Clinical characteristics of the studied sample.

| **Table 2. Clinical characteristics of the studied sample** |  |  |  |  |  |  |  |
| --- | --- | --- | --- | --- | --- | --- | --- |
|  | Healthy Controls (N=84) |  | Chronic Pain (N=157) |  | Statistics |  |  |
|  | Median | SD | Median | SD | Mann-Whitney U | z-statistics | p-values |
| PROMIS-SD8b total score | 51.70 | 8.39 | 58.30 | 8.11 | **9335.00** | **5.320** | **<0.001** |
| SF-36 (index) total score | 76.81 | 14.55 | 45.00 | 17.82 | **1745.00** | **-9.402** | **<0.001** |
| CTQ total score | 33.50 | 12.71 | 41.00 | 19.51 | **8497.50** | **3.694** | **<0.001** |
|  | Mean | SD | Mean | SD | t-value | df | p-value |
| BRS total score | 20.58 | 5.03 | 17.31 | 5.33 | **4.628** | **239** | **<0.001** |
| NRS – current | - | - | 3.80 | 2.60 | - | - | - |
| NRS – past week | - | - | 4.92 | 2.33 | - | - | - |
|  | N | % | N | % | X<sup>2</sup> | df | p-values |
| Mental health conditions (Yes) | 19 | 23% | 95 | 61% | **33.245** | **1** | **<0.001** |
| Depression | 11 | 13% | 69 | 44% |  |  |  |
| Anxiety disorder | 6 | 7% | 33 | 21% |  |  |  |
| Somatic symptom disorder | 0 | 0% | 17 | 11% |  |  |  |
| Eating Disorder | 1 | 1% | 9 | 6% |  |  |  |
| Obsessive compulsive disorder | 1 | 1% | 1 | 1% |  |  |  |
| Post-traumatic stress disorder | 2 | 2% | 30 | 19% |  |  |  |
| Personality disorder | 1 | 1% | 7 | 4% |  |  |  |
| Addiction disease | 0 | 0% | 5 | 3% |  |  |  |
| Adjustment disorder | 5 | 6% | 19 | 12% |  |  |  |
| Other | 5 | 6% | 8 | 5% |  |  |  |
| No information | 1 | 1% | 1 | 1% |  |  |  |
| More than one diagnosis | 7 | 8% | 51 | 32% |  |  |  |
| Physical health conditions (Yes) | 42 | 50% | 141 | 90% | **47.455** | **1** | **<0.001** |
| Asthma | 5 | 6% | 24 | 15% |  |  |  |
| Diabetes | 1 | 1% | 5 | 3% |  |  |  |
| Cancer | 0 | 0% | 7 | 4% |  |  |  |
| Chronic Obstructive Pulmonary Disease | 0 | 0% | 1 | 1% |  |  |  |
| Cardiovascular disease | 5 | 6% | 17 | 11% |  |  |  |
| Disease of the gastrointestinal tract | 1 | 1% | 25 | 16% |  |  |  |
| Hyper-/Hypothyroidism | 4 | 5% | 36 | 23% |  |  |  |
| Multiple sclerosis | 0 | 0% | 1 | 1% |  |  |  |
| Epilepsy | 2 | 2% | 1 | 1% |  |  |  |
| Other | 31 | 37% | 116 | 74% |  |  |  |
| More than one diagnosis | 6 | 7% | 57 | 36% |  |  |  |

|  |  |  |  |  |  |
| --- | --- | --- | --- | --- | --- |
| Pain conditions |  |  |  |  |  |
|  | Chronic backpain | - | - | 34 | 22% |
|  | Fibromyalgia | - | - | 36 | 23% |
|  | Migraine | - | - | 41 | 26% |
|  | Endometriosis | - | - | 30 | 19% |
|  | Arthritis | - | - | 14 | 9% |
|  | Irritable bowel syndrome | - | - | 9 | 6% |
|  | Neuropathic pain | - | - | 16 | 10% |
|  | Rheumatism | - | - | 22 | 14% |
|  | Gout | - | - | 1 | 1% |
|  | Osteoarthritis | - | - | 18 | 11% |
|  | Tension headache | - | - | 23 | 15% |
|  | Cluster headache | - | - | 3 | 2% |
|  | Crohn's disease | - | - | 0 | 0% |
|  | Ulcerative colitis | - | - | 3 | 2% |
|  | Lipoedema | - | - | 10 | 6% |
|  | Other | - | - | 37 | 24% |
|  | More than one diagnosis | - | - | 80 | 51% |
| Pain medications |  |  |  |  |  |
|  | Ibuprofen | - | - | 56 | 36% |
|  | Naproxen | - | - | 10 | 6% |
|  | Diclofenac | - | - | 7 | 4% |
|  | Paracetamol | - | - | 27 | 17% |
|  | Morphine | - | - | 5 | 3% |
|  | Oxycodone | - | - | 6 | 4% |
|  | Hydrocodone | - | - | 1 | 1% |
|  | Fentanyl | - | - | 53 | 34% |
|  | Buprenorphine | - | - | 53 | 34% |
|  | Hydromorphone | - | - | 3 | 2% |
|  | Tilidine | - | - | 14 | 9% |
|  | Novalgine | - | - | 48 | 31% |
|  | Tramadol | - | - | 6 | 4% |
|  | Amitriptyline | - | - | 9 | 6% |
|  | Imipramine | - | - | 53 | 34% |
|  | Clomipramine | - | - | 53 | 34% |
|  | Gabapentin | - | - | 4 | 3% |
|  | Pregabalin | - | - | 12 | 8% |
|  | Other | - | - | 40 | 25% |

|  |  |  |  |  |
| --- | --- | --- | --- | --- |
| No information | - | - | 53 | 34% |
| More than one painkiller | - | - | 76 | 48% |

SD: standard deviation; df: degrees of freedom; PROMIS-SD8b: PROMIS Sleep Disturbances 8b questionnaire; SF-36: 36 items short form survey; CTQ: childhood trauma questionnaire; BRS: brief resilience scale; NRS: numeric rating scale
Statistically significant tests are in bold

### Moderated mediation analysis

Statistical details of the moderated serial mediation are provided in Table 3 and represented Figure 2A.

**Figure 2.**
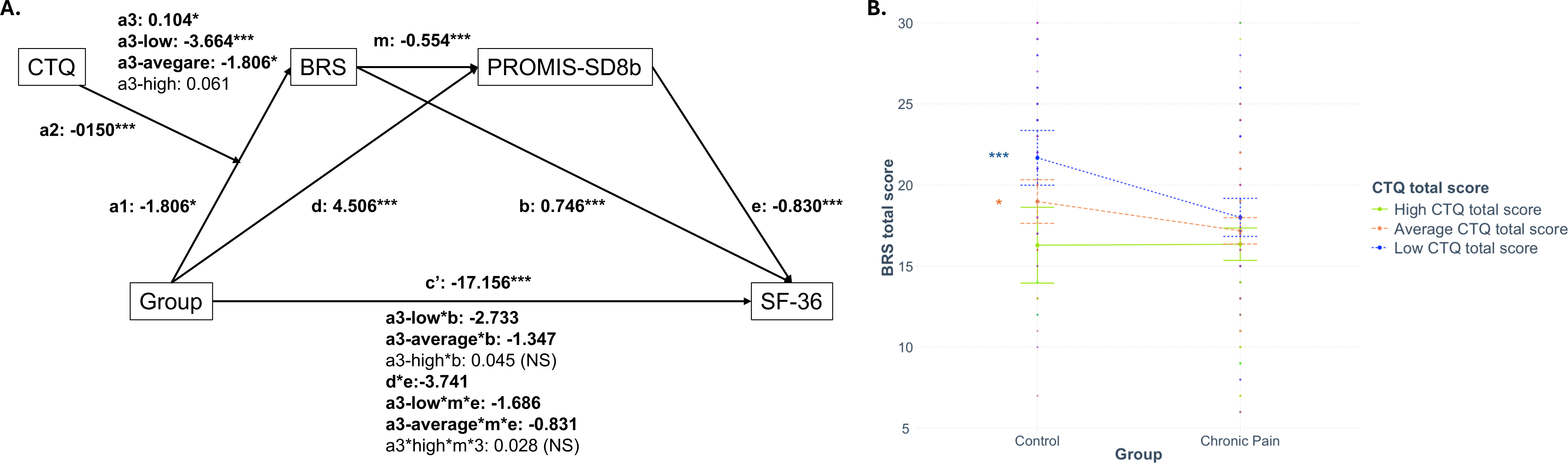
Illustration of the moderated serial mediation model tested. (A) Illustration and statistical details of the full moderated serial mediation model tested. (B) Graphic representation of the moderation of the group difference in resilience (BRS) by the severity of childhood maltreatment (CTQ total score). The levels of resilience were significantly higher in the control group at low (in blue dotted line) and average (orange dashed line), but not high (green plain lone) levels of childhood maltreatment. CTQ: childhood trauma questionnaire; BRS: brief resilience scale; PROMIS-SD8b: PROMIS Sleep Disturbances 8b questionnaire; SF-36: 36 items short form survey (quality of life). Statistically significant tests (*p*<0.05) are in bold. \**p*<0.05; \*\**p*<0.01; \*\*\**p*<0.001

**Table 3.**
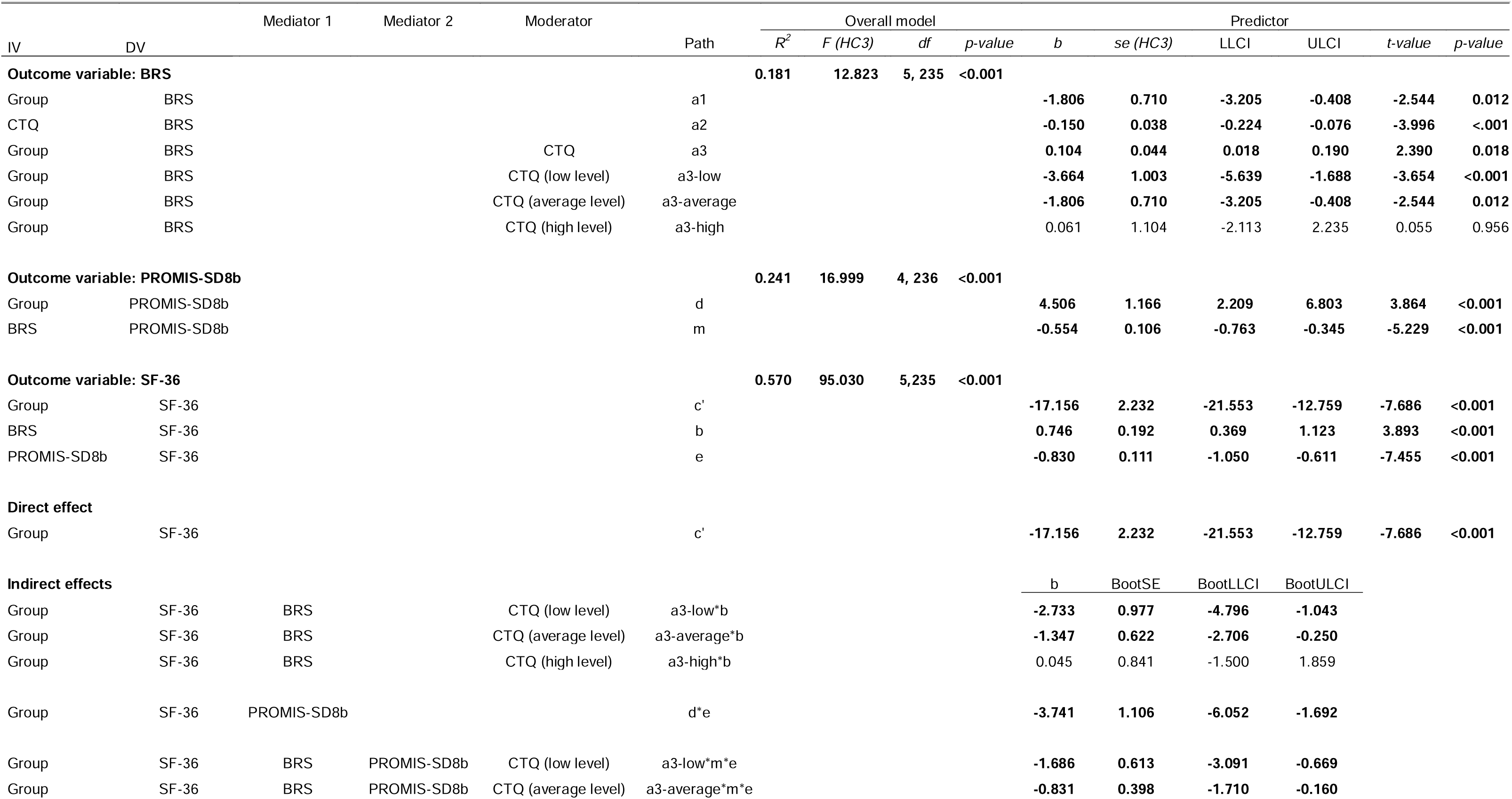
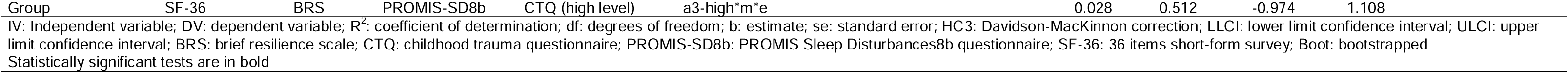
Statistical details of the moderated serial mediation model tested.

#### Moderated mediation 1

Levels of resilience were significantly lower in the chronic pain group relative to the control group (path a1) and associated with increasing childhood maltreatment severity (CTQ total score; path a2). The group-by-maltreatment interaction was significantly associated with variations in levels of resilience (path a3). Moderation analyses indicate that the group difference on levels of resilience was dependent of the severity childhood maltreatment. In particular, the chronic pain group reported lower levels of resilience compared to the control group at low (path a3-low), average (path a3-average) but not at high levels (path a3-high) of childhood maltreatment severity (see Figure 2B). In addition, lower quality of life was significantly associated with lower levels of resilience (path b1). The indirect effect of group on quality of life via resilience, moderated by childhood maltreatment, was only significant for people exposed to low (path a3-low*b1) and average (path a3-average*b1), but not high levels of childhood maltreatment (path a3-high*b1). In addition, quality of life was significantly lower in the chronic pain group compared to controls (direct path c’).

#### Mediation 2

Chronic pain was also significantly associated with greater sleep disturbances than controls (path d), and greater sleep disturbances were significantly associated with lower quality of life levels (path e). The indirect effect of group on quality of life, via sleep disturbances, was significant (path d*e).

#### Moderated serial mediation

The relationship between levels of resilience and sleep disturbances was significant (path m), indicating that lower levels of resilience were associated with greater sleep disturbances, independently of chronic pain. When both mediators are included in the same model, the indirect effect (path a*m*e) was significant and was further moderated by the severity of childhood maltreatment, with the indirect effects remaining significant for people exposed to low (path a3-low*m*e) and average (path a3-average*m*e), but not high (path a3-high*m*e) levels of childhood maltreatment.

## Discussion

The present study investigated the differences in the relationships between resilience and sleep disturbances on quality of life, existing between participants with chronic pain and controls, and whether the severity of childhood maltreatment moderates the relationship between group and resilience to impact sleep quality and quality of life. In line with previous research (Hunfeld, Perquin et al. 2001, Dysvik, Lindstrøm et al. 2004, Phyo, Freak-Poli et al. 2020), individuals with chronic pain reported significantly lower quality of life compared to controls. Beyond this, the moderated serial mediation model revealed that resilience and sleep disturbances jointly explained some of the group differences in quality of life, with childhood maltreatment moderating the group effect on resilience. Specifically, the group differences on quality of life through lower resilience, and sleep quality were stronger when participants reported at low and average, but not at high, levels of childhood maltreatment, indicating a ceiling effect of higher levels of childhood maltreatment on the group difference in resilience.

The model integrates and extends prior research by demonstrating that resilience and sleep disturbances do not operate in isolation, but together, to impact the quality of life of people experiencing chronic pain, relative to controls. Consistent with earlier findings (Morete, Solano et al. 2018, Chng, Yeo et al. 2023), individuals with chronic pain exhibited reduced resilience compared to controls. Importantly, this difference was most pronounced at low and average levels of childhood maltreatment exposure, supporting the notion that chronic pain and early adversity interact in shaping psychological resources. These findings align with studies reporting that childhood maltreatment is associated with lower resilience (Ellis, Bianchi et al. 2017, Nishimi, Choi et al. 2020) and extend them by showing that resilience may be particularly compromised when both chronic pain and a history of maltreatment co-occur, impacting the quality of life of people with chronic pain. The absence of group differences at high levels of maltreatment suggests that severe adversity may exert a ceiling effect on resilience, independently of experiencing chronic pain. This indicates a threshold at which early adversity uniformly diminishes resilience irrespective of experiencing chronic pain.

Resilience, in turn, was negatively associated with sleep disturbances, in line with evidence that adaptive coping skills and psychological flexibility buffer against sleep-related problems (Liu, Liu et al. 2016, Cai, Wang et al. 2021). Consistent with previous studies, elevated sleep disturbances were subsequently associated lower quality of life (Veldhuijzen, Greenspan et al. 2008, Rezaei, Mokhayeri et al. 2020). By integrating these components, the present model advances existing knowledge by identifying a conditional, sequential process: chronic pain is associated with lower levels of resilience particularly among those reporting exposure to lower levels of childhood maltreatment, thus increasing vulnerability to sleep disturbances, ultimately resulting in poorer quality of life. This relationship was not evident when people reported higher levels of childhood maltreatment, indicating a potential ceiling effect where people with chronic pain and those without show similar relationship to resilience when exposed to higher levels of childhood maltreatment. While separate associations between chronic pain and quality of life (Dysvik, Lindstrøm et al. 2004, Hadi, McHugh et al. 2019), sleep disturbances (Karaman, Karaman et al. 2014, Husak and Bair 2020), and resilience (Morete, Solano et al. 2018) have been established, these factors have not been investigated together in a comprehensive model. Moreover, although childhood maltreatment has been proposed as a risk factor for chronic pain (Davis, Luecken et al. 2005, Burke, Finn et al. 2017, Beal, Kashikar-Zuck et al. 2020), the present study is the first, to our knowledge, to determine its moderating role in the relationship between chronic pain and resilience. The current findings demonstrate that the interplay of childhood maltreatment, resilience, and sleep disturbances jointly accounts for lower quality of life in people with chronic pain. This integrative perspective underscores that psychosocial mechanisms should not be considered in isolation but as part of a complex and conditional system.

Taken together, these findings also highlight important implications for clinical practice. A more nuanced understanding of how resilience and sleep disturbances jointly contribute to reduced quality of life in chronic pain underscores the usefulness of interventions targeting both domains. Evidence-based (e.g., sleep hygiene training, cognitive-behavioural interventions for insomnia, or resilience-enhancing programs) and aetiology-informed interventions (e.g., trauma-focused cognitive behavioural therapies, eye movement desensitization and reprocessing or sensorimotor psychotherapy and somatic experiencing) may serve as promising avenues to improve daily functioning and well-being in individuals with chronic pain. Implementing such interventions in combination, particularly early after chronic pain develops, may help prevent further declines in quality of life by simultaneously strengthening psychological resources and reducing sleep-related vulnerability. By identifying resilience and sleep disturbances as modifiable mechanisms, the present study points to actionable targets for treatment and prevention aimed at improving long-term outcomes in people living with chronic pain.

In addition to strengths including a comprehensive assessment of the roles of childhood maltreatment, resilience, sleep disturbances in quality of life in chronic pain within the same model, this study also has several limitations. First, this study included a high proportion of females; this is not surprising as females more often report experiencing chronic pain (Rustøen, Wahl et al. 2004), and are more likely to participate in online studies (Wu, Zhao et al. 2022). In the overall sample, men make up about 20%, while in the chronic pain group, they account for only 10%. Since the general population has a more balanced gender distribution, this may limit the generalizability of the results. Second, the recruitment was made primarily through social media (e.g., Instagram), which mainly reaches younger adults. Promotion by influencers focusing on specific pain conditions, such as migraines, may have led to clustering of certain conditions. Third, 19% of the experimental group reported having chronic pain without a formal diagnosis, which is common in the general population. Fourth, the relatively high rate of comorbid mental health conditions (61% in the chronic pain group; 23% in the control group), although consistent with rates observed in the general population (14%) (World Health 2025), may also have influenced some of the relationships within the model. Fifth, this study was cross-sectional and associative in nature and does not provide causal interpretation of the relationship between the studied variables. Sixth, some participants from the control group reported acute bodily pain at the time of assessment (SF-36 item 21). Although this does not compare to pain experienced in chronic pain conditions, we cannot rule out the potential influence of acute pain on some of the variables of interest. Lastly, the sample had a high level of education, potentially limiting generalizability, and resilience was measured with only five items, raising questions about its adequacy in capturing such a broad construct.

In conclusion, the present study has identified that lower quality of life in people with chronic pain is dependent of individuals resilience abilities and sleep quality, and that the role of resilience on quality of life is dependent of the severity of childhood maltreatment reported. Maybe not surprising, results indicate that interventions that increase resilience and sleep quality may improve the quality of life for individuals with chronic pain. To strengthen these findings, replication of this study with a more balanced gender and pain condition distribution is needed. Additionally, analysing all quality of life subscales could clarify whether differences in quality of life are linked to specific aspects. Future research is warranted to better understand the causal relationship between these factors, to better identify targets for interventions and treatments across chronic pain conditions.

## Acknowledgments

The authors are grateful to the study participants for their time and participation.

## Data availability

The data that support the findings cannot be made publicly available due to ethical restrictions.

## Authors contributions

R.P. contributed conceptualization, data collection, methodology, validation, writing of the original draft, review and editing. F.S. contributed conceptualization, data collection, methodology, and review and editing. S.M.G. contributed conceptualization, data curation, formal analysis, investigation, methodology, project administration, resources, supervision, and review and editing. I.S. contributed conceptualization, data curation, formal analysis, investigation, methodology, project administration, resources, supervision, and review and editing. Y.Q. contributed conceptualization, data curation, formal analysis, investigation, methodology, validation, visualization, supervision, writing of the original draft, and review and editing.

## References

Arnow, B. A., E. M. Hunkeler, C. M. Blasey, J. Lee, M. J. Constantino, B. Fireman, H. C. Kraemer, R. Dea, R. Robinson and C. Hayward (2006). “Comorbid depression, chronic pain, and disability in primary care.” Psychosomatic Medicine 68(2): 262–268.

Arora, T., I. Grey, L. Östlundh, A. Alamoodi, O. M. Omar, K.-B. Hubert Lam and M. Grandner (2022). “A systematic review and meta-analysis to assess the relationship between sleep duration/quality, mental toughness and resilience amongst healthy individuals.” Sleep Medicine Reviews 62: 101593.

Bader, K., C. Hänny, V. Schäfer, A. Neuckel and C. Kuhl (2009). “Childhood Trauma Questionnaire – Psychometrische Eigenschaften einer deutschsprachigen Version.” Zeitschrift für Klinische Psychologie und Psychotherapie.

Beal, S. J., S. Kashikar-Zuck, C. King, W. Black, J. Barnes and J. G. Noll (2020). “Heightened risk of pain in young adult women with a history of childhood maltreatment: a prospective longitudinal study.” PAIN 161(1): 156.

Bernstein, D. P., J. A. Stein, M. D. Newcomb, E. Walker, D. Pogge, T. Ahluvalia, J. Stokes, L. Handelsman, M. Medrano, D. Desmond and W. Zule (2003). “Development and validation of a brief screening version of the Childhood Trauma Questionnaire.” Child Abuse & Neglect 27(2): 169–190.

Breivik, H., B. Collett, V. Ventafridda, R. Cohen and D. Gallacher (2006). “Survey of chronic pain in Europe: prevalence, impact on daily life, and treatment.” European Journal of Pain (London, England) 10(4): 287–333.

Bullinger, M., I. Kirchberger and J. Ware (1995). “Der deutsche SF-36 Health Survey Übersetzung und psychometrische Testung eines krankheitsübergreifenden Instruments zur Erfassung der gesundheitsbezogenen Lebensqualität.” Zeitschrift für Gesundheitswissenschaften = Journal of public health 3(1): 21–36.

Burke, A. L. J., J. L. Mathias and L. A. Denson (2015). “Psychological functioning of people living with chronic pain: a meta-analytic review.” The British Journal of Clinical Psychology 54(3): 345–360.

Burke, N. N., D. P. Finn, B. E. McGuire and M. Roche (2017). “Psychological stress in early life as a predisposing factor for the development of chronic pain: Clinical and preclinical evidence and neurobiological mechanisms.” Journal of Neuroscience Research 95(6): 1257–1270.

Buysse, D. J., L. Yu, D. E. Moul, A. Germain, A. Stover, N. E. Dodds, K. L. Johnston, M. A. Shablesky-Cade and P. A. Pilkonis (2010). “Development and Validation of Patient-Reported Outcome Measures for Sleep Disturbance and Sleep-Related Impairments.” Sleep 33(6): 781–792.

Cai, Y., J. Wang and L. Hou (2021). “Resilience Improves the Sleep Quality in Disabled Elders: The Role of Perceived Stress.” Frontiers in Psychology 12.

Carpi, M., C. Cianfarani and A. Vestri (2022). “Sleep Quality and Its Associations with Physical and Mental Health-Related Quality of Life among University Students: A Cross-Sectional Study.” International Journal of Environmental Research and Public Health 19(5): 2874.

Chmitorz, A., M. Wenzel, R.-D. Stieglitz, A. Kunzler, C. Bagusat, I. Helmreich, A. Gerlicher, M. Kampa, T. Kubiak, R. Kalisch, K. Lieb and O. Tüscher (2018). “Population-based validation of a German version of the Brief Resilience Scale.” PLoS ONE 13(2): e0192761.

Chng, Z., J. J. Yeo and A. Joshi (2023). “Resilience as a protective factor in face of pain symptomatology, disability and psychological outcomes in adult chronic pain populations: a scoping review.” Scandinavian Journal of Pain 23(2): 228–250.

Cohen, J., P. Cohen, S. G. West and L. S. Aiken (2003). Applied multiple regression/correlation analysis for the behavioral sciences, 3rd ed. Mahwah, NJ, US, Lawrence Erlbaum Associates Publishers.

Cohen, S. P., L. Vase and W. M. Hooten (2021). “Chronic pain: an update on burden, best practices, and new advances.” Lancet (London, England) 397(10289): 2082–2097.

Davis, D. A., L. J. Luecken and A. J. Zautra (2005). “Are reports of childhood abuse related to the experience of chronic pain in adulthood? A meta-analytic review of the literature.” The Clinical Journal of Pain 21(5): 398–405.

Davison, A. C. and D. V. Hinkley (1997). Bootstrap Methods and their Application. Cambridge, Cambridge University Press.

Dillmann, U., P. Nilges, H. Saile and H. U. Gerbershagen (2011). “PDI - Pain Disability Index - deutsche Fassung.”

Ding, H., J. Han, M. Zhang, K. Wang, J. Gong and S. Yang (2017). “Moderating and mediating effects of resilience between childhood trauma and depressive symptoms in Chinese children.” Journal of Affective Disorders 211: 130–135.

Dysvik, E., T. C. Lindstrøm, O.-J. Eikeland and G. K. Natvig (2004). “Health-related quality of life and pain beliefs among people suffering from chronic pain.” Pain Management Nursing: Official Journal of the American Society of Pain Management Nurses 5(2): 66–74.

Edwards, R. R., R. H. Dworkin, M. D. Sullivan, D. C. Turk and A. D. Wasan (2016). “The Role of Psychosocial Processes in the Development and Maintenance of Chronic Pain.” The Journal of Pain 17(9 Suppl): T70–92.

Ellis, B. J., J. Bianchi, V. Griskevicius and W. E. Frankenhuis (2017). “Beyond Risk and Protective Factors: An Adaptation-Based Approach to Resilience.” Perspectives on Psychological Science: A Journal of the Association for Psychological Science 12(4): 561–587.

Faul, F., E. Erdfelder, A. Buchner and A.-G. Lang (2009). “Statistical power analyses using G*Power 3.1: Tests for correlation and regression analyses.” Behavior Research Methods 41(4): 1149–1160.

Fritz, J., A. M. de Graaff, H. Caisley, A.-L. van Harmelen and P. O. Wilkinson (2018). “A Systematic Review of Amenable Resilience Factors That Moderate and/or Mediate the Relationship Between Childhood Adversity and Mental Health in Young People.” Frontiers in Psychiatry 9.

Gerbershagen, H. U., G. Lindena, J. Korb and S. Kramer (2002). “[Health-related quality of life in patients with chronic pain].” Schmerz (Berlin, Germany) 16(4): 271–284.

Greger, H., A. Myhre, S. Lydersen and T. Jozefiak (2016). “Child maltreatment and quality of life: A study of adolescents in residential care.” Health and Quality of Life Outcomes 14.

Hadi, M. A., G. A. McHugh and S. J. Closs (2019). “Impact of Chronic Pain on Patients’ Quality of Life: A Comparative Mixed-Methods Study.” Journal of Patient Experience 6(2): 133–141.

Hayes, A. F. (2022). “Introduction to Mediation, Moderation, and Conditional Process Analysis: Third Edition: A Regression-Based Approach.” Guilford Press.

Hayes, A. F. and L. Cai (2007). “Using heteroskedasticity-consistent standard error estimators in OLS regression: An introduction and software implementation.” Behavior Research Methods 39(4): 709–722.

Hunfeld, J. A., C. W. Perquin, H. J. Duivenvoorden, A. A. Hazebroek-Kampschreur, J. Passchier, L. W. van Suijlekom-Smit and J. C. van der Wouden (2001). “Chronic pain and its impact on quality of life in adolescents and their families.” Journal of Pediatric Psychology 26(3): 145–153.

Husak, A. J. and M. J. Bair (2020). “Chronic Pain and Sleep Disturbances: A Pragmatic Review of Their Relationships, Comorbidities, and Treatments.” Pain Medicine 21(6): 1142–1152.

Karaman, S., T. Karaman, S. Dogru, Y. Onder, R. Citil, Y. E. Bulut, H. Tapar, A. Sahin, S. Arici, Z. Kaya and M. Suren (2014). “Prevalence of sleep disturbance in chronic pain.” European Review for Medical and Pharmacological Sciences 18(17): 2475–2481.

Karimi, M. and J. Brazier (2016). “Health, Health-Related Quality of Life, and Quality of Life: What is the Difference?” PharmacoEconomics 34(7): 645–649.

Kascakova, N., J. Furstova, R. Trnka, J. Hasto, A. M. Geckova and P. Tavel (2022). “Subjective perception of life stress events affects long-term pain: the role of resilience.” BMC Psychology 10(1): 54.

Kieselbach, K., M. Schiltenwolf and C. Bozzaro (2016). “Versorgung chronischer Schmerzen: Wirklichkeit und Anspruch.” Der Schmerz 30.

Leiner (2024). “SoSci Survey (Version 3.5.02).”

Liu, X., C. Liu, X. Tian, G. Zou, G. Li, L. Kong and P. Li (2016). “Associations of Perceived Stress, Resilience and Social Support with Sleep Disturbance Among Community-dwelling Adults.” Stress and Health 32(5): 578–586.

Morete, M. C., J. P. C. Solano, M. S. Boff, W. J. Filho and H. A. Ashmawi (2018). “Resilience, depression, and quality of life in elderly individuals with chronic pain followed up in an outpatient clinic in the city of São Paulo, Brazil.” Journal of Pain Research 11: 2561–2566.

Morfeld, M., I. Kirchberger and M. Bullinger (2011). “SF-36 - Fragebogen zum Gesundheitszustand | Testzentrale.”

Nicola, M., H. Correia, G. Ditchburn and P. Drummond (2021). “Invalidation of chronic pain: a thematic analysis of pain narratives.” Disability and Rehabilitation 43(6): 861–869.

Nishimi, K., K. W. Choi, K. A. Davis, A. Powers, B. Bradley and E. C. Dunn (2020). “Features of Childhood Maltreatment and Resilience Capacity in Adulthood: Results from a Large Community-Based Sample.” Journal of Traumatic Stress 33(5): 665–676.

Pagels, L., K. Lüdtke and A. Schäfer (2022). “Kinesiophobie bei Schulterbeschwerden: Validierung der deutschen Version der Tampa Scale for Kinesiophobia (TSK-GV).” Schmerz.

Park, J. Y., C. W. Lee, Y. Jang, W. Lee, H. Yu, J. Yoon, S. Oh, Y. S. Park, H. A. Ryoo, J. Lee, N. Cho, C. H. Lee, Y. C. Lee, H.-H. Won, H. S. Kang, T. H. Ha and W. Myung (2023). “Relationship between childhood trauma and resilience in patients with mood disorders.” Journal of Affective Disorders 323: 162–170.

Phyo, A. Z. Z., R. Freak-Poli, H. Craig, D. Gasevic, N. P. Stocks, D. A. Gonzalez-Chica and J. Ryan (2020). “Quality of life and mortality in the general population: a systematic review and meta-analysis.” BMC public health 20(1): 1596.

Purabdollah, M., S. Lakdizaji, A. Rahmani, M. Hajalilu and K. Ansarin (2015). “Relationship between Sleep Disorders, Pain and Quality of Life in Patients with Rheumatoid Arthritis.” Journal of Caring Sciences 4(3): 233–241.

Radbruch, L., G. Loick, P. Kiencke, G. Lindena, R. Sabatowski, S. Grond, K. A. Lehmann and C. S. Cleeland (1999). “Validation of the German version of the Brief Pain Inventory.” Journal of Pain and Symptom Management 18(3): 180–187.

Reimer, M. A. and W. W. Flemons (2003). “Quality of life in sleep disorders.” Sleep Medicine Reviews 7(4): 335–349.

Rezaei, O., Y. Mokhayeri, J. Haroni, M. J. Rastani, M. Sayadnasiri, H. Ghisvand, M. Noroozi and B. Armoon (2020). “Association between sleep quality and quality of life among students: a cross sectional study.” International Journal of Adolescent Medicine and Health 32(2).

Rustøen, T., A. K. Wahl, B. R. Hanestad, A. Lerdal, S. Paul and C. Miaskowski (2004). “Gender differences in chronic pain—findings from a population-based study of Norwegian adults.” Pain Management Nursing 5(3): 105–117.

Smith, B. W., J. Dalen, K. Wiggins, E. Tooley, P. Christopher and J. Bernard (2008). “The brief resilience scale: Assessing the ability to bounce back.” International Journal of Behavioral Medicine 15(3): 194–200.

Studer, M., J. Stewart, N. Egloff, E. Zürcher, R. von Känel, J. Brodbeck and M. grosse Holtforth (2017). “Psychosoziale Stressoren und Schmerzempfindlichkeit bei chronischer Schmerzstörung mit somatischen und psychischen Faktoren (F45.41).” Der Schmerz 31(1): 40–46.

Teoli, D. and A. Bhardwaj (2024). Quality Of Life. StatPearls. Treasure Island (FL), StatPearls Publishing.

Treede, R.-D., W. Rief, A. Barke, Q. Aziz, M. I. Bennett, R. Benoliel, M. Cohen, S. Evers, N. B. Finnerup, M. B. First, M. A. Giamberardino, S. Kaasa, E. Kosek, P. Lavand’homme, M. Nicholas, S. Perrot, J. Scholz, S. Schug, B. H. Smith, P. Svensson, J. W. S. Vlaeyen and S.-J. Wang (2015). “A classification of chronic pain for ICD-11.” Pain 156(6): 1003–1007.

Turk, D. C. and K. V. Patel (2022). Epidemiology and economics of chronic and recurrent pain. Clinical Pain Management, John Wiley & Sons, Ltd: 6–24.

Veldhuijzen, D. S., J. D. Greenspan and M. T. Smith (2008). Sleep and Quality of Life in Chronic Pain. Sleep and Quality of Life in Clinical Medicine. J. C. Verster, S. R. Pandi-Perumal and D. L. Streiner. Totowa, NJ, Humana Press: 187–197.

Weber, S., A. Jud and M. A. Landolt (2016). “Quality of life in maltreated children and adult survivors of child maltreatment: a systematic review.” Quality of Life Research 25(2): 237–255.

World Health, O. (2025). World mental health today: latest data, World Health Organization.

Wörz, R., J. Horlemann and G. H. H. Müller-Schwefe (2022). “Auswirkungen, Chronifizierung, Epidemiologie, zeitgemäße Diagnostik.” Schmerzmedizin 38(4): 46–50.

Wu, M.-J., K. Zhao and F. Fils-Aime (2022). “Response rates of online surveys in published research: A meta-analysis.” Computers in Human Behavior Reports 7: 100206.

Yazdi-Ravandi, S., Z. Taslimi, H. Saberi, J. Shams, S. Osanlo, G. Nori and A. Haghparast (2013). “The Role of Resilience and Age on Quality of life in Patients with Pain Disorders.” Basic and Clinical Neuroscience 4(1): 24–30.

Yu, L., D. J. Buysse, A. Germain, D. E. Moul, A. Stover, N. E. Dodds, K. L. Johnston and P. A. Pilkonis (2011). “Development of short forms from the PROMIS™ sleep disturbance and Sleep-Related Impairment item banks.” Behavioral Sleep Medicine 10(1): 6–24.

Yu, L., D. J. Buysse, A. Germain, D. E. Moul, A. Stover, N. E. Dodds, K. L. Johnston and P. A. Pilkonis (2012). “Development of Short Forms From the PROMIS™ Sleep Disturbance and Sleep-Related Impairment Item Banks.” Behavioral Sleep Medicine 10(1): 6–24.

